# Model-based projections for COVID-19 outbreak size and student-days lost to closure in Ontario childcare centers and primary schools

**DOI:** 10.1101/2020.08.07.20170407

**Authors:** Brendon Phillips, Dillon T. Browne, Madhur Anand, Chris T. Bauch

## Abstract

There is a pressing need for evidence-based scrutiny of plans to re-open childcare during the COVID-19 pandemic. Here we developed an agent-based model of SARS-CoV-2 transmission within a childcare center and households. Scenarios varied the student-to-educator ratio (15:2, 8:2, 7:3), and family clustering (siblings together vs. random assignment). We also evaluated a primary school setting (30:1, 15:1 and 8:1) including cohorts that alternate weekly. In the childcare scenarios, grouping siblings significantly reduced outbreak size and student-days lost. We identify an intensification cascade specific to classroom outbreaks of respiratory viruses with presymptomatic infection. In both childcare and primary school settings, each doubling of class size from 8 to 15 to 30 more than doubled the outbreak size and student-days lost, by factors of 2-5, respectively 2.5-4.5, depending on the scenario. Proposals for childcare and primary school reopening could be enhanced for safety by switching to lower ratios and sibling groupings.

## Introduction

As nations around the world grapple with the psychosocial, civic, and economic ramifications of social distancing guidelines, the critical need for widely-available Early Childhood Education (or colloquially, “childcare”) services have, once again, reached the top of policy agendas^1,2^. Whether arguments are centered on human capital (i.e., “children benefit from high-quality, licensed educational environments, and have the right to access such care”) or the economy (i.e., “parents need childcare in order to work, and the economy needs workers to thrive”), the conclusion is largely the same: childcare centers are re-opening, at least in some capacity, and this is taking place before a vaccine or herd immunity can mitigate potential spread of SARS-CoV-2 (the virus that causes COVID-19). Outbreaks of COVID-19 in emergency childcare centers and schools have already been observed^3^, causing great concern as governments struggle to balance “flattening the curve” and preventing second waves with other pandemic-related sequelae, such as the mental well-being of children and families, access to education and economic disruption.

Governments and childcare providers are tirelessly planning the operations of centers, with great efforts to follow public health guidelines for reducing SARS-CoV-2 contagion^4^. However, these guidelines, which will result in significantly altered operational configurations of childcare centers and substantial cost increases, have yet to be rigorously examined. Moreover, discussions of childcare are presently eclipsed by general discussion of “school” reopening^5^. That being said, for many parents, the viability of the school-day emerges from before and after school programming that ensures adequate coverage throughout parents’ work schedules. Yet, reopening plans often fail to mention the critical interplay between school and childcare, even though many childcare centers operate within local schools^6^. Consequently, a model that comprehensively examines the multifaceted considerations surrounding childcare operations may help inform policy and planning. As such, the purpose of the present investigation is to develop an agent-based model that explores and elucidates the multiple interacting factors that could impact potential SARS-CoV-2 spread in school-based childcare centers.

In Ontario, Canada (the authors’ jurisdiction), childcare centers were permitted to reopen on June 12, 2020, provided centers limit groupings (e.g., classrooms) to a maximum of 10 individuals (educators and children, inclusive)^7^. Additionally, all centers had to come up with a plan for daily screening of incoming persons, thorough cleaning of rooms before and during operations, removal of toys that pose risk of spreading germs, allowing only essential visitors, physical distancing at pick-up and drop-off, and a contingency plan for responding should anyone be exposed to the virus (e.g., closing a classroom or center for a period of time). Further school-specific recommendations have been recently outlined by The Toronto Hospital for Sick Children^6^, which include specific guidelines for screening, hand hygiene, physical distancing, cleaning, ventilation, and masking. While this influential report has become the guiding framework for school reopening in Ontario, there remains no discussion of childcare operations in relation to SARS-CoV-2 spread. Guidelines for primary schools call for either full re-opening, with up to 30 students per classroom attending every day, or with cohorts of 15 students attending in alternate weeks.

Simulation models of infectious disease spread have been widely applied during the COVID-19 pandemic, as in previous pandemics^8,9^. Modelling is used to determine how quickly the pathogen can spread^10^, how easily it may be contained^11^, and the relative effectiveness of different containment strategies^12,13^. Sensitivity analysis is crucial to assess whether model predictions are robust to uncertainties in data^14^, which is particularly important during a pandemic caused by a novel emerging pathogen like SARS-CoV-2. Agent-based models are particularly well-suited to situations where a highly granular description of the population is desirable and where random effects (stochasticity) is important. Such models have been previously applied in both pandemic and non-pandemic situations^15–17^, and is our choice of modelling methodology in the present work focusing on SARS-CoV-2 transmission in schools and households. Our objective was to use our agent-based model to project the impact of student-to-educator (or in the case of childcare centres, child-to-educator) ratios and sibling grouping strategies on outbreaks of COVID-19 and student-days lost to classroom closure in a hypothetical childcare center and primary school.

Below, the modelling approach, results, and interpretation of the present modelling exercise are described. In the following Methods section, the rationale and parameterization of the model are specified in detail. In the Results section, the performance of the model under different assumptions is showcased. We start with analyzing the childcare center setting and end with the primary school setting. Lastly, the discussion will provide a review and interpretation of this study, including any limitations and future suggestions for research.

## Model Overview

A detailed description of the model structure, assumptions and parameterization appears in the Methods section. We developed an agent-based model of SARS-CoV-2 transmission in a population structured into households and classrooms, as might represent a childcare setting or a small primary school (Figure 1A). Individuals were categorized into either child or adult, and contacts between these groups were parameterized based on contact matrices estimated for the Canadian setting. Household sizes were determined from Canadian demographic data. Classroom sizes and student-educator ratios were determined according to the scenario being studied. For the childcare setting we analyzed student-educator ratios of 8:2 and 7:3, giving a maximum class size of 10 representative of the smaller enrollment at schools. We also analyzed a student-educator ratio 15:2, giving a total class size of 17. Along with class size, we also consider class composition. Individuals may spread the infection to their household members each day, so effective contacts and interaction in the classroom may result in qualitatively different spreading patterns. As such, children in this model can be assigned to classrooms either randomly (*RA*) or by grouping siblings (or otherwise cohabiting students) together (ST) in an attempt to reduce SARS-CoV-2 transmission. For the primary school setting, we considered student-educator ratios of 8:1, 15:1, and 30:1, all with the random allocation. For the 8:1 and 15:1 ratios we also considered scenarios where cohorts of 8 or 15 students attending the same classroom but in alternating weeks. These scenarios were labelled 8(A):1 and 15(A):1. In the primary school setting, we considered the higher student-educator ratio 30:1 as an example of larger class size. Some plans considered in reopening Ontario educational institutions divides this larger class size into two alternating cohorts of 15 students each with a single shared educator; we call this scenario 15(A):1. Rotation occurs each week, so that one cohort engages with online material while the other receives face-to-face instruction for 5 days, after which the cohorts exchange roles. The student-educator ratios 8:1 and 8(A):1 were also included for comparison to smaller class sizes. For primary schools we considered only the RA allocation.

**Figure 1.**
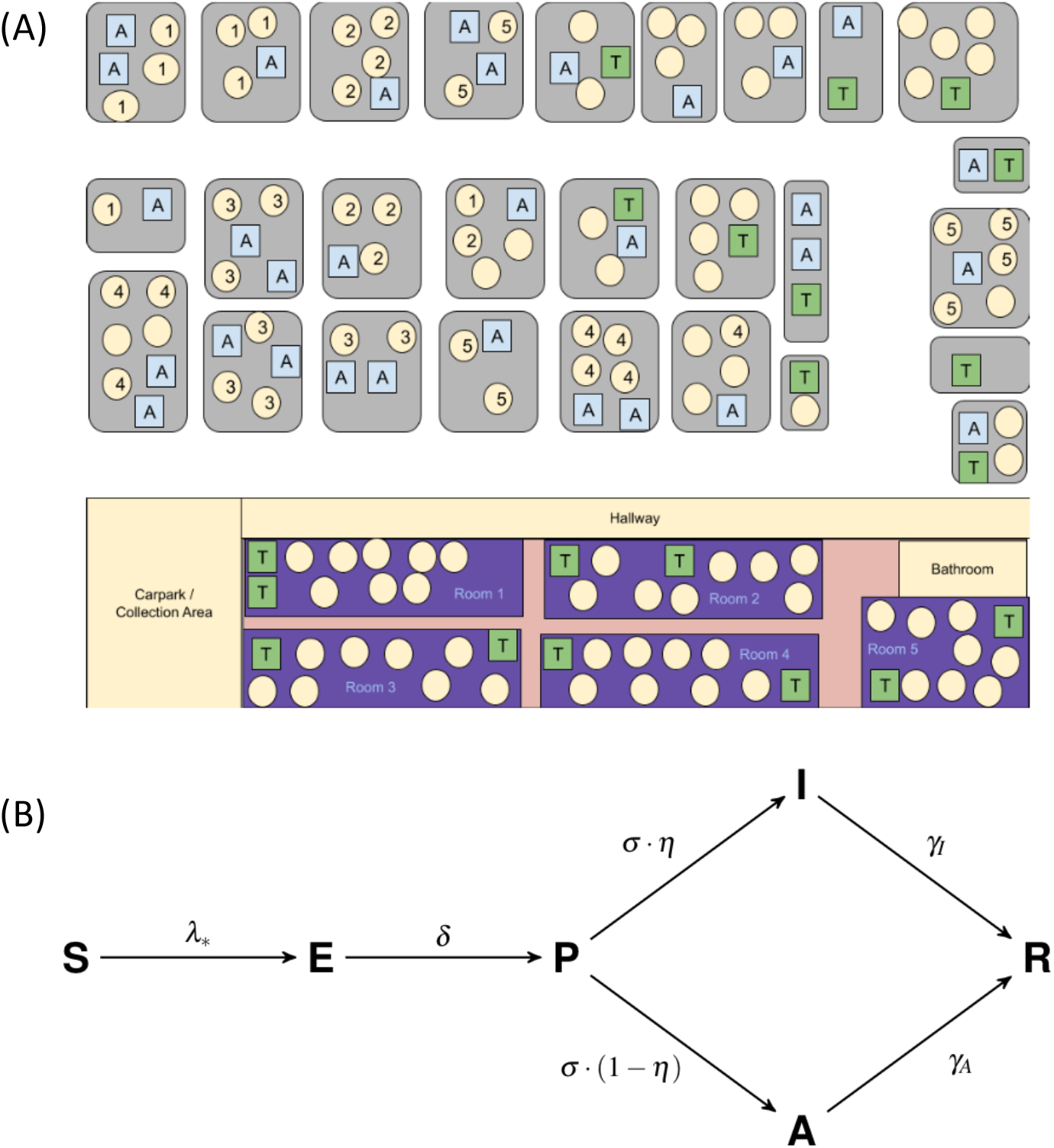
(A) Schematic representation of model population. ‘A’ represents adult, ‘T’ represents educator, and circles represent children. Grey rectangles represent houses and the school is represented at the bottom of the figure. Numbers exemplify possible assignments of children in households to classrooms. (B) Diagram showing the SEPAIR infection progression for each agent in the simulation (see Methods for definitions of parameters).

SARS-CoV-2 could be transmitted in households, classrooms or in common areas of the school, all of which were treated as homogeneously mixing on account of evidence for aerosolized routes of transmission^18^. Individuals were also subject to a constant background risk of infection from other sources, such as shopping centers. Figure 1B shows the progression of the illness experienced by each individual in the model. In each day, susceptible (*S*) individuals exposed to the disease via community spread or interaction with infectious individuals (those with disease statuses *P*, *A* and *I*) become exposed (*E*), while previously exposed agents become presymptomatic (*P*) with probability *δ*. Presymptomatic agents develop an infection in each day with probability *δ*, where they can either become symptomatically infected (*I*) with probability n or asymptomatically infected (*A*) with probability 1 − *η*. If a symptomatic individuals appears in a classroom, that classroom is closed for 14 days (in the case of alternating cohorts for primary schools, we assumed both cohorts are closed). Other classrooms in the same school may remain open. Asymptomatic students and educators return at the end of this period while symptomatic students and educators remain at home and symptomatic educators are replaced by substitutes.

Children are less affected by the SARS-CoV-2 virus than adults, and account for a smaller proportion of COVID-19 cases^19^. However, the role of children in SARS-CoV-2 transmission is still debated, and existing epidemiological evidence is limited by lack of empirical studies in school settings, which have been closed for much of 2020. Other studies show that children shed a similar amount of virus to adults^20^. To account for this ambiguity, we used contact matrices drawn from populations under ‘business as usual’ circumstances as a proxy of what contact rates would look like under a full reopening of schools and workplaces^21^, but but we considered both a high transmission rate scenario and a low transmission rate scenario. The low transmission rate scenario represented either reduced transmission rates in children, and/or highly effective infection control through consistent use of high-effectiveness masks, social distancing, and disinfection protocols (see Methods section for details). In total the permutations on student-educator ratios, transmission rate assumptions, siblings versus non-sibling groupings, and alternating cohorts yielded 22 scenarios (Table 1).

**Table 1.**
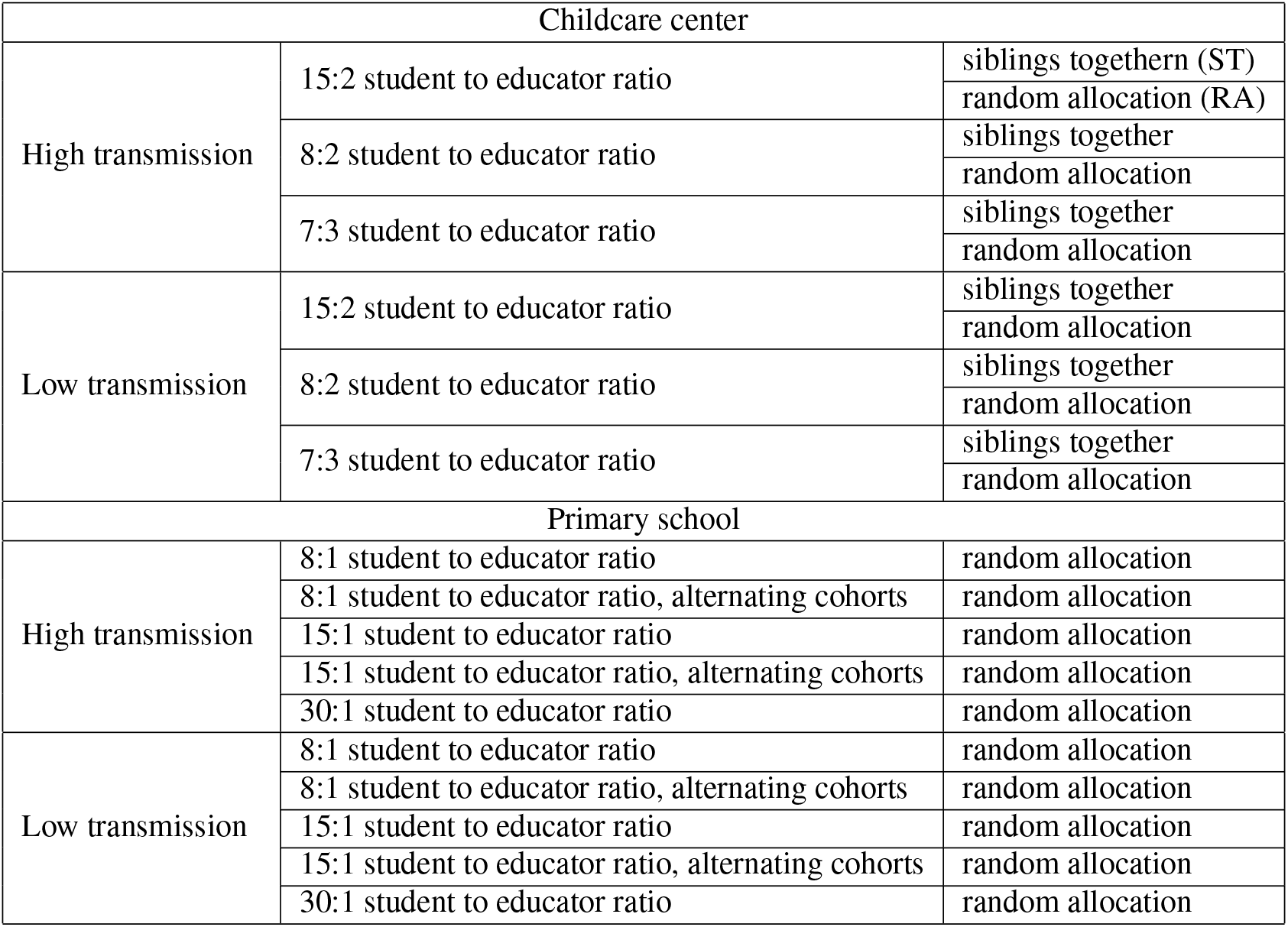
22 Scenarios evaluated based on different assumptions about transmission probabilities, educator-student ratios, and student allocation.

## Results

### Initial stages of the outbreak

The time evolution of the outbreaks are illustrated in Fig. 2, which shows the proportion of actively infected school attendees (both children and educators) per day in twelve childcare center scenarios. Many of the scenarios tend to produce a well-defined outbreak curve close to the start of the simulation, even with classroom closure protocols in place. However, the outbreaks are more strongly household-driven for the 7:3 and 8:2 ratios than the 15:2 ratio; this is apparent in the weekly waves superimposed on the overall epidemic curve more strongly in the 15:2 scenarios, on account of the impact of weekends. The 15:2 ratio also tends to generate earlier, more intense outbreaks, while 7:3 and 8:2 scenarios produce fewer infections that are more sporadically distributed throughout the simulated time horizon. In the case of high transmission, the maximum mean level of exposure (*E*) is 4.97% in the 15:2 RA configuration 18 days into the the simulation, on average, with peak 3.03% presymptomatic (*P*) and 1.64% asymptomatic (*A*) attendees at days 12 and 19 respectively. Meanwhile, peak mean exposure in scenario 7:3 ST occurs on day 2, with 1.9% attendees exposed to the disease, with presymptomatic cases never exceeding that of the start of any simulation.

**Figure 2.**
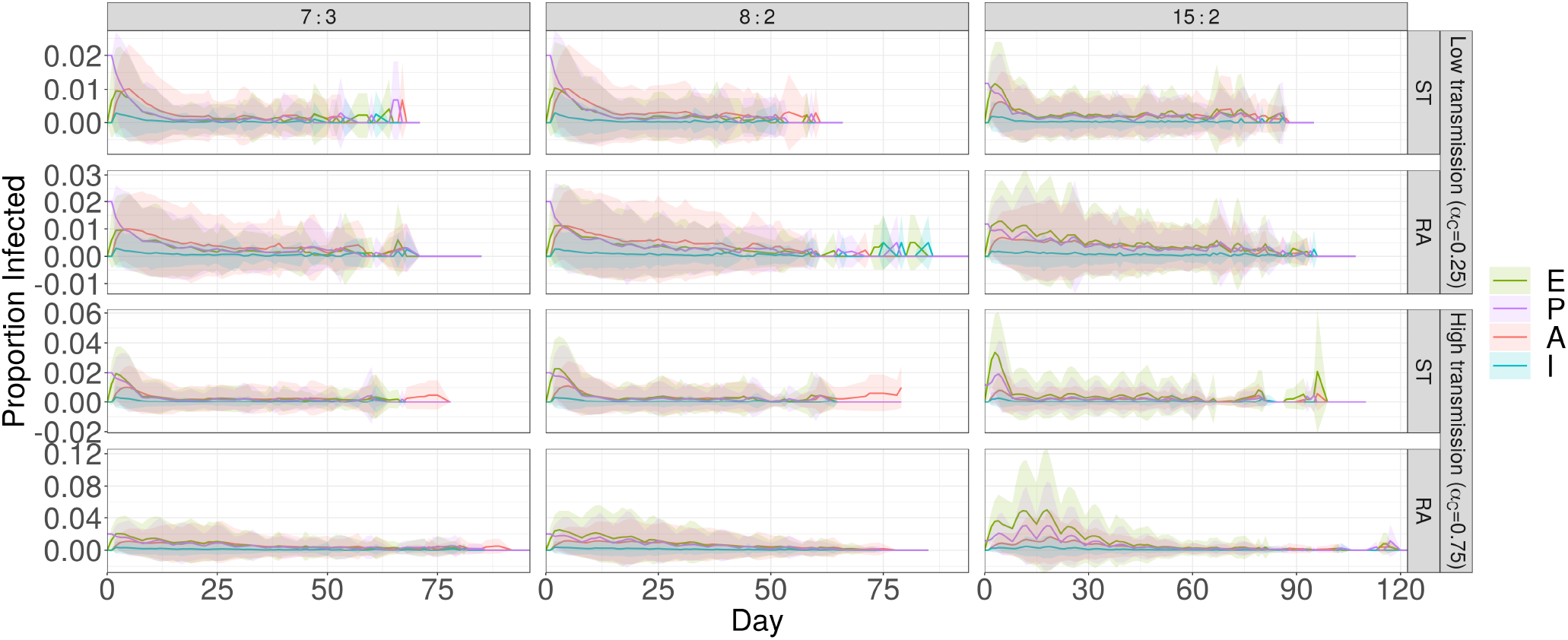
Time series of the proportions of exposed (*E*), presymptomatic (*P*), asymptomatic (*A*) and infected (*I*) individuals in the simulation for each scenario. The ensemble means are represented by solid lines, while the respected shaded ribbons show one standard deviation of the results.

Supplementary Tab. S1 summarizes the information from the figures, showing the days until the 30-day peak of each proportion of active infections in the center. Here we can see that active infections peak far earlier with the ST allocation than with the RA allocation for both high (*α =* 0.75) and low (*α* = 0.25) transmission rates in most cases, and have either equal or smaller peaks for most maximum proportions corresponding to the RA allocation independent of student-educator ratio. In the case of high transmission, peak proportions decrease with the number of students per class in half of the tested scenarios (statuses *P* and *I* with RA allocation, and status *E*). In the low transmission case, there is a reversal in trend, with peak proportions increasing with decreasing number of exposed (*E*) and presymptomatic (*P*) students per class. There is no obvious relationship between peak days for infected (*I*) and asymptomatic (*A*) individuals in the high transmission case, neither for asymptomatic (*A*) individuals in the low transmission case.

The basic reproduction number *R*_0_ is the average number of secondary infections produced by a single infected person in an otherwise susceptible population^22^. When there is pre-existing immunity, as we suppose here, we study the effective reproduction number *R_e_* - the average number of secondary infections produced by a single infected person in a population with some pre-existing immunity. Supplementary Fig. S1A shows the estimated *R_e_* and mean population size (school plus all associated households) over the course of each simulation, computed by tracking the number of secondary infections produced by a single primary case. The *R_e_* values measured from the simulation range from 1.5 to 3 on average, depending on the scenario. These *R_e_* values are generally lower than the typical range of *R*_0_ values between 2 to 3 reported in the literature^23^. This is the expected relationship, not only because of pre-existing immunity, but also because the *R_e_* values in our simulation capture transmission only in schools and workplaces, while the *R*_0_ values in the literature are measured for SARS-CoV-2 transmission in all settings, including workplaces and other sources of community spread.

There is little correlation between mean population size (Supplementary Fig. S1A, line), number of households (not shown) and the corresponding *R_e_* estimate (Supplementary Fig. S1A, bars), leaving only the number of children per classroom responsible for the gross increasing trend in *R_e_* in both high (*α* = 0.75) and low (*α* = 0.25) transmission scenarios. Equation 3 shows that child-child contact within the classroom occurs at least 2 times more often than any other type of contact; given that the majority of the attendees of the school are children, we can expect *R_e_* to depend on the number of children enrolled in the school.

This is further demonstrated by the bar charts of Supplementary Fig. S1B, which show the distribution of times between the primary infection case and the first secondary infection. The scenarios with the highest ratio of children to educators (15:2) show the quickest start of the outbreak in both high and low transmission cases, with RA having the highest proportion of trials where the first secondary infection occurred within a single day in the high transmission case. In comparison, scenario 7:3 RA showed the slowest average initial spread in the high transmission case, while the low transmission case sees low rates for both 8:2 and 7:3. Configuration ST (except for ratio 7:3) frequently results in faster secondary spread over the first two days (even in the first 2 weeks).

### Outbreak duration

Each individual simulation end when all classes are at full capacity and there are no active infections in the population-aside from community infection, this marks the momentary halt of SARS-CoV-2 spread. From this, we get a description of the duration of the first outbreak. (There could well be a second outbreak sparked by some community infection among individuals who remain susceptible at the end of the first outbreak). Box plots in Fig. 3 show that the 15:2 ratio in both RA and ST allocations gives a median outbreak duration at least as large as all other scenarios (for both low and high transmission cases). Another general observation is that classroom allocation (RA vs. ST) doesn’t change the distribution of outbreak duration for student-educator ratios 8:2 and 7:3 as drastically as it does for 15:2, whereas ST allocation results in lower median duration (24 vs 43 for RA allocation) and significantly lower maximum duration for the 15:2 ratio (61 vs. 88 for RA allocation without outliers) in the high transmission case.

**Figure 3.**
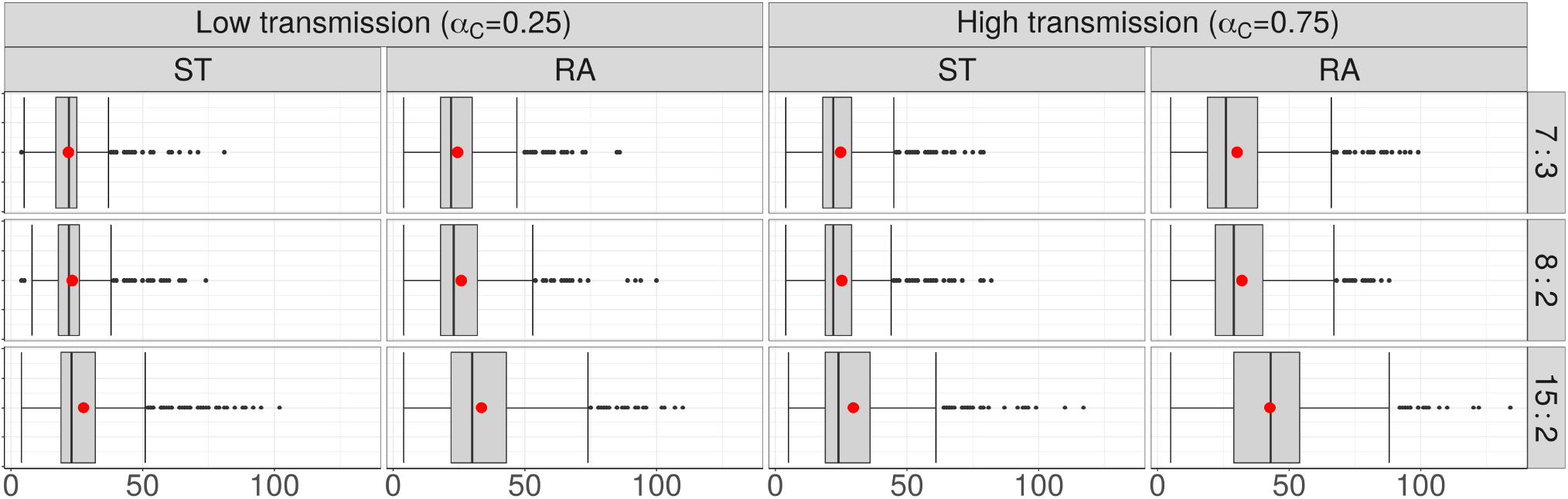
Box plots depicting the distribution of simulation durations for each scenario. Taken together with the stopping criteria of the simulations and measures of aggregate, these describe the duration of the outbreak. Red dots represent the arithmetic mean of the data.

This is mirrored in the low transmission case as well. A possible explanation lies in the number of students per classroom. The child-child contact rate (Eqn. 3) is far higher than any other contact rate, implying that the classroom is the site of greatest infection spread (demonstrated in Fig. 4A). ST allocation differs from RA allocation in its containment of disease transfer from the classroom to a comparatively limited number of households. This effect (the difference between ST and RA) is amplified with the addition of each new student to the classroom, so that while the difference between 7:3 and 8:2 may be small (only 1 student added), the effect becomes far exaggerated when the student number is effectively doubled (15 students vs. 7 or 8).

**Figure 4.**
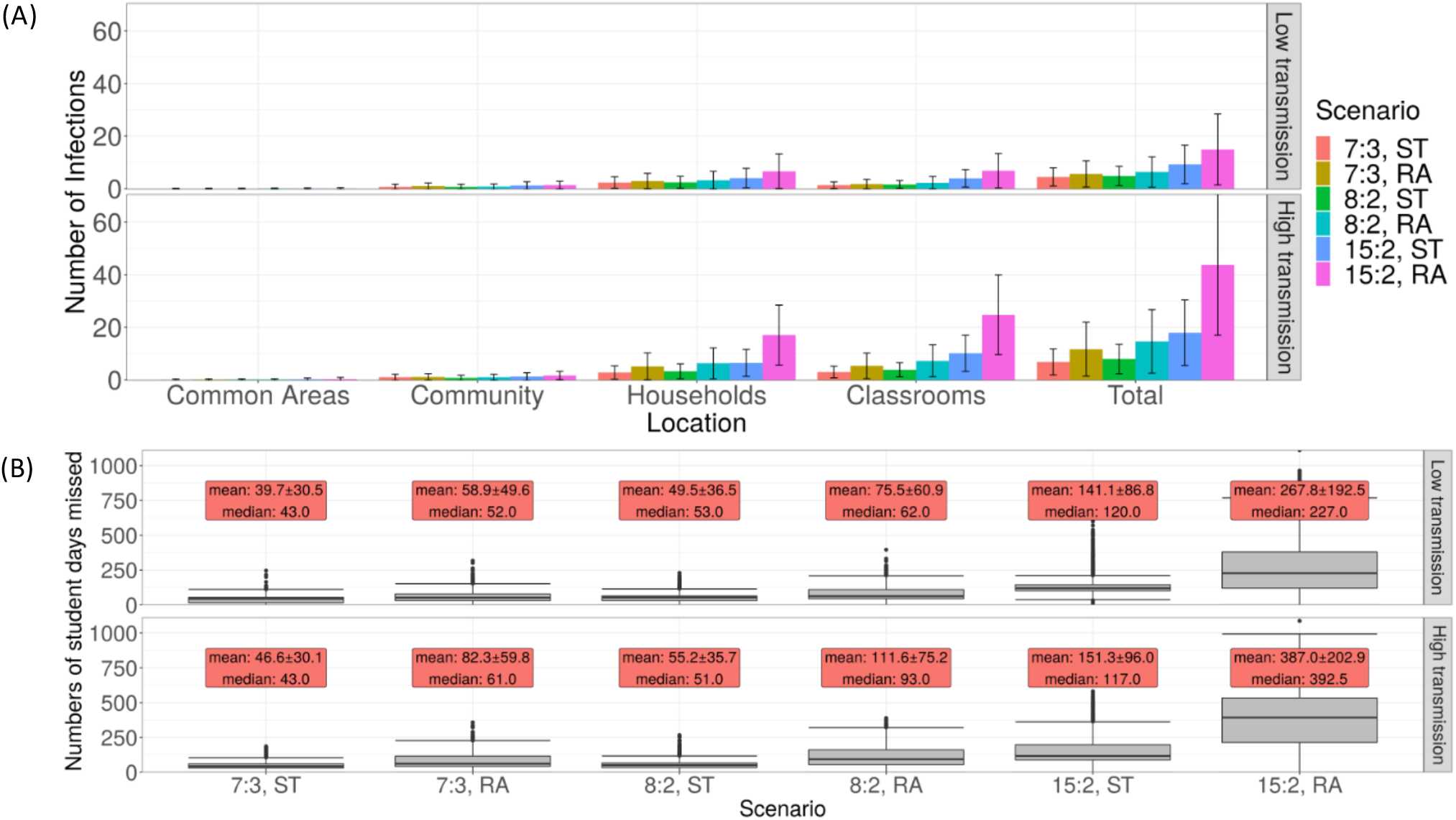
COVID-19 outbreak size and student-days lost to closure in the childcare setting. (A) the mean number of infections occurring among all school attendees in each location over time for each scenario. The height of each bar gives the ensemble mean and its standard deviation is represented by error bars. (B) Box plots showing the number of student days forfeited over the course of the simulation due to class closure upon the detection of an outbreak. Red text boxes show the mean and standard deviation of closure.

The evolution of the numbers of susceptible (*S*) and recovered/removed (*R*) school attendees provides additional information on the course of the outbreak, since they represent the terminal states of the infection process in each individual by the end of the outbreak. Supplementary Fig. S1C shows the proportion of susceptible and recovered current school attendees (who have not been sent home due to classroom outbreaks). As with all results so far, the 15:2 RA scenario most efficiently facilitates disease spread through the school in both high and low transmission cases, with the proportion of recovered attendees (*R*) overtaking the number of never-infected attendees (status *S*) on day 34 in the case of high transmission (*α* = 0.75). Performance between 8:2 and 7:3 with ST allocation is similar for both transmission rates, though all scenarios show smaller variation over trials featuring lower infection transmission. As shown in Fig. 3, scenario 15:2 RA gave the longest average simulation time in the high transmission scenario; this is also reflected in Supplementary Fig. 2, where the longest outbreak lasted 134 days.

### Outbreak size and classroom closure

Figure 4A shows the mean number of infections in each location in all scenarios, as well as the total number of infections in each scenario (the ‘outbreak size’). As expected, many more infections occur in the high transmission scenario (*α* = 0.75), and the error bars of the plot show greater standard deviation of the results than in the low transmission (*α* = 0.25) scenario. But for each location and regardless of the transmission rate scenario, the number of infections increases rapidly with the number of children in the classroom in each room allocation. The 15:2 ratio is universally the worst allocation across all possible scenarios. However, the difference between the outbreak size in different scenarios decreases as the transmissibility of the virus drops (so to speak, the gap been between the 15:2 RA and 15:2 ST scenarios decreases as *α* decreases, and so with other student-educator ratios). When the transmission rate is high, the relatively larger variety (by household) and prevalence of child-child interactions has a multiplicative effect on the number of effective transmissions in the classroom. Lower transmissibility thereby decreases the classroom infection rates relative to the household transmission rates.

The numbers of student-days forfeited due to classroom closure are given in Fig. 4B, according to scenario. (The number of student-days forfeited is the number of days of closure times the number of students who would otherwise have been able to continue attending.) In all scenarios, the 15:2 student-educator ratio is quantitatively the worst strategy examined by almost an order of magnitude, resulting in the highest possible number of student-days forfeited. RA allocation shows worse performance than ST in all scenarios. Both the low (*α* = 0.25) and high (*α* = 0.75) transmissibility scenarios favour the 7:3 student-educator ratio and ST allocation, with a lower number of student-days forfeited. The poor performance of 15:2 ratio occurs because it suffers from a multiplicative effect: larger class sizes are more likely to be the origin of outbreak, and when the outbreak starts, more children are affected when the classroom is shut down. Moreover, since it’s possible for a student or educator to be infected during a 14-day closure, not all attendees necessarily return to class upon reopening; sick educators are replaced with substitutes. As such, these class closures results in otherwise healthy students missing potentially additional school days beyond the 14-day closure period. The 15:2 strategy suffers particularly from this effect, since transmission is facilitated when more students are in a classroom.

Naturally, a high incidence of COVID-19 cases will result in multiple room closures; one way to see this is to look at the number and duration of room closures, both shown in Fig. S1D. In all scenarios, schools spent (on average) more days with one closed classroom than any other number. We can also observe a difference in RA and ST allocations for the 7:3 ratio: with both high and low transmission rate (*α* = 0.25 and *α* = 0.75 respectively), RA allocation results in a higher number of class closures.

### Primary school settings

The primary school setting shows the same cascade of intensifying outbreaks and rapidly mounting student-days of closure as class sizes increase (Fig. 5). This effect occurs in both childcare centres and primary schools because firstly, in a larger classroom it is more likely that a student tests positive for COVID-19. Secondly, when the classroom closes as a result, more students are affected by the closure. Thirdly, because COVID-19 is characterized by presymptomatic infection and aerosol dispersal, there is more infection in larger classrooms before the closure is enacted. Introducing more children into the classroom increases the effective reproductive ratio (*R_e_*) for both low and high rates of transmission while cohorting/alternation has little effect (Supplementary Fig. S2A), and similar strategies (that is, differing by only 1 student or educator per class, or by alternation) give similar reproductive ratios *R_e_* (compare to Supplementary Fig. S1A).

**Figure 5.**
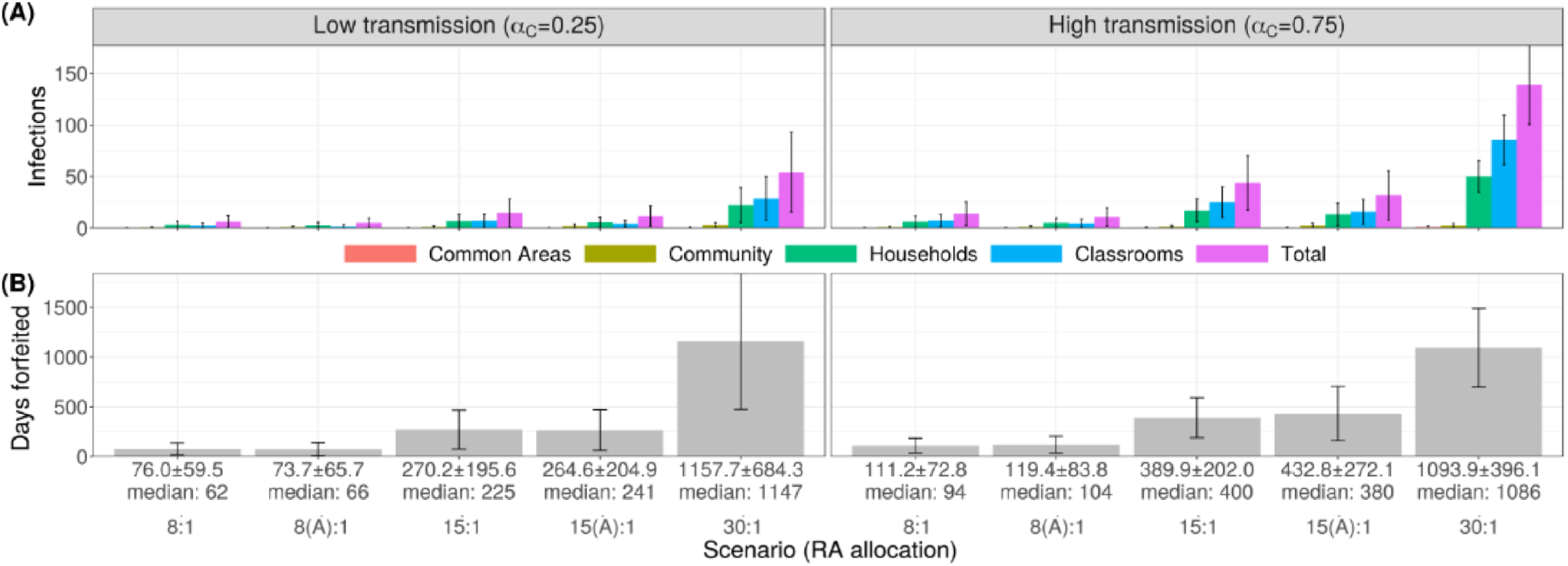
COVID-19 outbreak size and student-days lost to closure in the childcare setting. (A) Bar charts showing the mean number of infections occurring in each location over the time of the simulation, (B) Bar charts showing the number of student days forfeited due to classroom closures sparked by disease outbreak. Error bars represent one standard deviation of the corresponding data.

There is little difference between numbers of forfeited student days between the similar scenarios 8:1 and 8(A):1, as well as as 15:1 and 15(A):1 (Fig. 5B). Since the shutdown of a classroom affects both cohorts, there will be very little difference in virus spread between scenarios allotting the same number of students per classroom. This effect is also seen in Supplementary Fig. 5A. Comparison of Fig. 4A and Fig. 5A show similar distributions of outbreak size for all student-teacher ratios, signifying that cohorting does not significantly change the results of structured interactions featured in the model. The true benefit of cohorting arises in the consideration of class sizes, given the desire for contact time with all enrolled students. Comparison of Fig. 4B and Fig. 5B shows that the similar scenarios 15:2 RA, 15:1 RA and 15(A):1 RA all result in a comparable number of forfeited student-days in both low and high transmission scenarios, as do the scenarios 8:2 RA, 8:1 RA and 8(A):1 RA.

Higher student-educator ratios facilitate faster disease spread through the school than smaller ones (Supplementary Fig. S2B). One major difference is the weekly fluctuation of the infection status curves visible in the cohorted scenarios 8(A):1 and 15(A):1. These fluctuations correspond to the rotation of the student cohorts through the school term. Transitions between majority susceptible and recovered regimes is delayed (high transmission) or prevented (low transmission) by cohorting; we see that alternating strategies result in better aggregate infection outcomes, even when classroom capacity is held constant. Scenario 15(A):1 also results in shorter mean and median outbreak lengths in the entire population in both low and high transmission cases (Supplementary Fig. S2C).

### Sensitivity Analysis

We conducted a sensitivity analysis on *β^H^*, *β^C^*, *λ* and *R_init_* (see Supplementary Appendix for details). We found that variation in rates of household and classroom interaction and infection (*β^H^* and *β^C^*) and the number of individuals initially recovered (*R_init_*) greatly impact SARS-CoV-2 transmission, but did not change the relative performances of the 22 scenarios. The greatest influence on outcomes remain the scheme of allocation of students to classrooms (RA or ST), the number of students per class (15, 8 or 7), and whether the transmission rate in the classrooms is low or high (*α_C_*). Other important factors include classroom closure upon identification of a symptomatic case and the interaction patterns of asymptomatic infected individuals in the household upon classroom closure (i.e. whether they continue to interact in close contact, as would be necessary for younger children, or whether children are old enough to effectively self-isolate). Our baseline assumption was to assume asymptomatic infected individuals who are sent home due to closure of a classroom are able to self-isolate. This assumption is conservative, since inability to self-isolate under these circumstances would result in higher projected outbreak sizes.

## Discussion

We developed and simulated an agent-based model of SARS-CoV-2 transmission in childcare center and primary school settings for the purposes of informing reopening policies. The model was configured to capture SARS-CoV-2 transmission in a local school building, since many childcare centers operate across several classrooms within schools. These services are an essential bridge for many parents who are unable to drop-off or pick-up children around school hours due to work. Our findings suggest that variability in class size (i.e., number of children in a class) and class composition (i.e., sibling groupings versus random assignment) influence the nature of SARS-CoV-2 transmission within the childcare context. Specifically, a 7:3 student-to-educator ratio that utilized sibling groupings yielded the lowest rates of transmission, while a 15:2 ratio consistently performed far worse. Findings for the primary school ratios show a similar acceleration of negative impacts with increasing class size. Findings from our simulations are sobering, as educators in the province lobbied for a 15 student cap on classrooms in Summer 2020. Our study suggests that classes of this size pose a tangible risk for COVID-19 outbreaks, and that lower ratios would better offset infection and school closures. While school reopening guidelines^6^, public health agencies^24^, and public petitions^25^ have called for smaller class sizes, governments appear to be following some recommendations in reopening plans while ignoring others.

This accelerating effect of increasing classroom sizes occurs because of three factors working in concert. Firstly, a larger class means that a student is more likely to test positive for COVID-19 at some point. Secondly, when a larger class is closed as a result, it affects more students. Third, presymptomatic transmission and higher densities of students ensure that more children become infected before classroom closure is enacted, resulting in larger outbreak sizes due to more cases both before the closure, and after the closure as the infection continues to spread in households. This particular mechanism is specific to institutional outbreaks for infectious diseases with pre-symptomatic transmission worsened by aerosol transmission routes^18^.

Policies related to childcare and traditional school reopening have not been well integrated^26^. In Ontario, childcare classrooms were capped at a maximum of 10 occupants, overall (hence the 8:2 and 7:3 ratios in the present study)^7^. Conversely, procedures for traditional “school” classrooms have been given the go-ahead for 15 children (hence the 15:2 ratio). While allowable class sizes will differ somewhat as a function of child age and jurisdiction, it seems likely that early childhood and elementary school classes may actually surpass these numbers in Ontario. Our findings demonstrate that the 15:2 ratio represents a significantly higher risk, not only for SARS-CoV-2 spread, but for school closures. In one scenario (15:2 random assignment), the modeled outbreak lasted for 105 days. Given that childcare and schools are often operating within the same physical location, this policy discrepancy is questionable. Based on our simulations, a lower ratio (7:3) is indicated. Moreover, it appears that this configuration could be enhanced through the utilization of sibling groupings.

An examination of student days missed due to classroom closure further elucidates the favorability of smaller class size and sibling grouping as a preventative measure. In this analysis, the worst configuration was the 15:2 random assignment ratio. Again, this was observed in both high transmission and low transmission environments. In the most unfavorable scenario (15:2 RA), there were cumulatively 387 and 267 student days forfeited in high versus low transmission settings, respectively. Conversely, in the best scenario (7:3, siblings together), there were only 47 and 40 student days forfeited. Thus, our simulations suggest that the lower ratios and sibling groupings offer a safeguard against high disruptive classroom closures^27,28^. Given this, a proactive and preventative approach that builds in realistic levels of reduced class time would be better than a reactive strategy that yields unpredictable closure events due to outbreaks.

Several policy and procedural recommendations have emerged from this modeling exercise. First, it is recommended that childcare and school settings, alike, consider lowering student-to-educator ratios. Commensurate with the present findings, a 7:3 ratio (10 individuals per class including both children and adults) outperforms a 15:2 ratio on key metrics. Second, there also appears to be benefit associated with sibling groupings. Thus, a siblings together configuration should be considered. Third, the majority of transmission occurred in the classroom. As such, it is important for reopening plans to consider social distancing and hygiene procedures within classrooms - a recommendation that may only be feasible with fewer children in the classroom. It is unlikely that classrooms with 15 or more children will afford children with the necessary space to socially distance. Finally, in the primary school settings, significant benefits accrue for 15(A):1 relative to the 30:1 student-educator ratio, and thus decision-makers should reconsider the conventional model of putting 30 students in classrooms every day in favour of cohorts of 15 students alternating weekly.

Finally, the present study has a number of limitations that should be considered. While it is becoming increasingly clear that COVID-19 risk varies as a function of social determinants of health (e.g., socioeconomic status, race, ethnicity, immigration status, neighborhood risk), along with opportunities for social distancing^29^, the present study did not take these considerations into account. Future simulation studies might consider how these social determinants intersect with childcare and school configurations. Additionally, this study was primarily concerned with SARS-CoV-2 infection and student days lost. That being said, there are many important outcomes to consider in relation to children’s developmental health in the pandemic. Longitudinal studies considering children’s learning and mental health outcomes in relation to new childcare and school configurations are strongly indicated^30^.

## Materials and Methods

### Population Structure

There are *N* households in the population, and a single educational institution (either a school or a school, dependent on scenarios to be introduced later) with *M* rooms and a maximum capacity dependent on the scenario being tested. Effective contacts between individuals occur within each household, as well as rooms and common areas (entrances, bathrooms, hallways, etc.) of the institution. All groups of individuals (households and rooms) in the model are assumed to be well-mixed.

Each individual (agent) in the model is assigned an age, household, room in the childcare facility and an epidemiological status. Age is categorical, so that every individual is either considered a child (C) or an adult (A). Epidemiological status is divided into stages in the progression of the disease; agents can either be susceptible (*S*), exposed to the disease (*E*), presymptomatic (an initial asymptomatic infections period *P*), symptomatically infected (*I*), asymptomatically infected (*A*) or removed/recovered (*R*), as shown in Fig. 1B.

In the model, some children in the poFpulation are enrolled as students in the institution and assigned a classroom based on assumed scenarios of classroom occupancy while some adults are assigned educator/caretaker roles in these classroom (again dependent on the occupancy scenario being tested). Allocations are made such that there is only one educator per household and that children do not attend the same institution as a educator in the household (if there is one), and *vice versa*.

### Interaction and Disease Progression

The basic unit of time of the model is a single day, over which each attendee (of the institution) spends time at both home and at the institution. The first interactions of each day are established within each household, where all members of the household interact with each other. An asymptomatically infectious individual of age *i* will transmit the disease to a susceptible housemate with the age *j* with probability 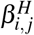, while symptomatically infectious members will self-isolate (not interact with housemates) for a period of 14 days.

The second set of interpersonal interactions occur within the institution. Individuals (both students and educators) in each room interact with each other, where an infectious individual of age *i* transmits the disease to some susceptible individual of age *j* with probability 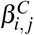. To signify common areas within the building (such as hallways, bathrooms and entrances), each individual will then interact with every other individual in the institution. There, an infectious individual of age *j* will infect a susceptible individual of age *i* with probability 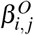.

To simulate community transmission (for example, public transport, coffee shops and other sources of infection not explicitly modelled here), each susceptible attendee is infected with probability *λ_S_*. Susceptible individuals not attending the institution in some capacity are infected at rate *λ_N_*, where *λ_N_* > *λ_S_* to compensate for those consistent effective interactions outside of the institution that are neglected by the model (such as workplace interactions among essential workers and members of the public).

Figure 1B shows the progression of the illness experienced by each individual in the model. In each day, susceptible (*S*) individuals exposed to the disease via community spread or interaction with infectious individuals (those with disease statuses *P*, *A* and *I*) become exposed (*E*), while previously exposed agents become presymptomatic (*P*) with probability *δ*. Presymptomatic agents develop an infection in each day with probability *δ*, where they can either become symptomatically infected (*I*) with probability *η* or asymptomatically infected (*A*) with probability 1 − *η*.

The capacity of the sole educational institution in the model is divided evenly between 5 rooms, with class size and student-educator ratio governed by one of three basic scenarios: seven students and three educators per room (7: 3), eight students and two educators per room (8: 2), and fifteen students and two educators per room (15: 2). Classroom allocations for children can be either randomised or grouped by household (siblings are put in the same class).

Symptomatically infected agents (*I*) are removed from the simulation after 1 day (status *R*) with probability *γ_I_*, upon which they self-isolate for 14 days, and therefore no longer pose a risk to susceptible individuals. Asymptomatically infected agents (*A*) remain infectious but are presumed able to maintain regular effective contact with other individuals in the population due to their lack of noticeable symptoms; they recover during this period (status R) with probability *γ_A_*. Disease statuses are updated at the end of each day, after which the cycles of interaction and infection reoccur the next day.

The actions of symptomatic (status *I*) agents depend on age and role. Individuals that become symptomatic maintain a regular schedule for 1 day following initial infection (including effective interaction within the institution, if attending), after which they serve a mandatory 14-day isolation period at home during which they interaction with no one (including other members of their household). On the second day after the individual’s development of symptoms, their infection is considered a disease outbreak centerd in their assigned room, triggering the closure of that room for 14 days. All individuals assigned to that room are sent home, where they self-isolate for 14 days due to presumed exposure to the disease. Symptomatically infected children are not replaced, and simply return to their assigned classroom upon recovery. At the time of classroom reopening, any symptomatic educator is replaced by a substitute for the duration of their recovery, upon which they reprise their previous role in the institution; the selection of a substitute is made under previous constraints on educator selection (one educator per household. with no one chosen from households hosting any children currently enrolled in the institution).

### Parameterization

The parameter values are given in Supplementary Tab. S2. The sizes of households in the simulation was determined from 2016 Statistics Canada census data on the distribution of family sizes^31^. We note that Statistics Canada data only report family sizes of 1, 2 or 3 children: the relative proportions for 3+ children were obtained by assuming that 65% of families of 3+ children had 3 children, 25% had 4 children, 10% had 5 children, and none had more than 5 children. Each educator was assumed to be a member of a household that did not have children attending the school. Again using census data, we assumed that 36% of educators live in homes with no children, where an individual lives alone with probability 0.282, while households hosting 3, 4, 5, 6, and seven adults occur with probability 0.345, 0.152, 0.138, 0.055, 0.021 and 0.009 respectively. Others live with ≥ 1 children in households following the size and composition distribution depending on the number of adults in the household. For single-parent households, a household with a single child occurs with probability 0.169, and households with 2, 3, 4 and 5 children occur with probabilities 0.079, 0.019, 0.007 and 0.003 respectively. With two-parent households, those probabilities become 0.284, 0.307, 0.086, 0.033 and 0.012.

The age-specific transmission rates in households are given by the matrix:

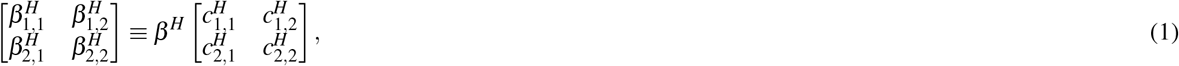

where 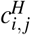 gives the number of contacts per day reported between individuals of ages *i* and *j* estimated from data^21^ and the baseline transmission rate *β^H^* is calibrated. To estimate 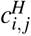 from the data in Ref.^21^, we used the non-physical contacts of age class 0-9 years and 25-44 years of age with themselves and one another in Canadian households. Based on a meta-analysis, the secondary attack rate of SARS-CoV-2 appears to be approximately 15% on average in both Asian and Western households^32^. Hence, we calibrated *β^H^* such that a given susceptible person had a 15% chance of being infected by a single infected person in their own household over the duration of their infection averaged across all scenarios tested (App.). As such, age specifictransmission is given by the matrix

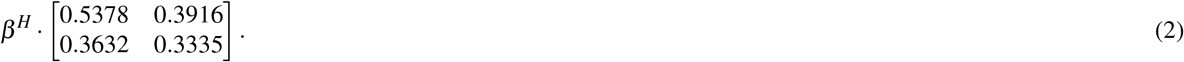

To determine *λ_S_* we used case notification data from Ontario during lockdown, when schools, workplaces, and schools were closed^33^. During this period, Ontario reported approximately 200 cases per day. The Ontario population size is 14.6 million, so this corresponds to a daily infection probability of 1.37 × 10^−5^ per person. However, cases are under-ascertained by a significant factor in many countries^34^–we assumed an under-ascertainment factor of 8.45, meaning there are actually 8.45 times more cases than reported in Ontario, giving rise to *λ_S_ =* 1.16 × 10^−4^ per day; *λ_N_* was set to 2 · *λ_S_*.

The age-specific transmission rates in the school rooms is given by the matrixPCi

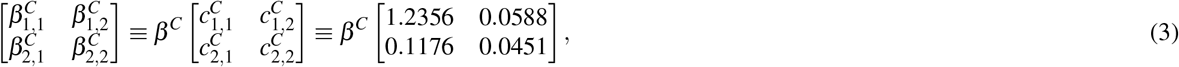

where 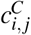 is the number of contacts per day reported between age *i* and *j* estimated from data^21^. To estimate 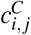 from the data in Ref.^21^, we used the non-physical contacts of age class 0-9 years and 20-54 years of age, with themselves and one another, in Canadian schools. Epidemiological data on secondary attack rates in childcare settings are rare, since schools and schools were closed early in the outbreak in most areas. We note that contacts in families are qualitatively similar in nature and duration to contacts in schools with small group sizes, although we contacts are generally more dispersed among the larger groups in rooms, than among the smaller groups in households. On the other hand, rooms may represent equally favourable conditions for aerosol transmission, as opposed to close contact. Hence, we assumed that *β^C^ = α_C_β^H^*, with a baseline value of *α_C_ =* 0.75 based on more dispersed contacts expected in the larger room group, although we varied this assumption in sensitivity analysis.

To determine *β^O^* we assumed that *β^O^ = α_O_β^C^* where *α_O_* ≪ 1 to account for the fact that students spend less time in common areas than in their rooms. To estimate *α_O_*, we note that *β^O^* is the probability that a given infected person transmits the infection to a given susceptible person. If students and staff have a probability *p* per hour of visiting a common area, then their chance of meeting a given other student/staff in the same area in that area is *p*^2^. We assumed that *p =* 0.05 and thus *α_O_ =* 0.0025. The age-specific contact matrix for *β^O^* was the same as that used for *β^C^* (Eqn. 3).

### Model Initialisation

Upon population generation, each agent is initially susceptible (*S*). Individuals are assigned to households as described in the Parameterisation section, and children are assigned to rooms either randomly or by household. We assume that parents in households with more than one child will decide to enroll their children in the same institution for convenience with probability *ξ* = 80%, so that each additional child in multi-child households will have probability 1 − *ξ* of not being assigned to the institution being modelled.

Households hosting educators are generated separately. As in the Parameterisation section, we assume that 36% of educators live in adult-only houses, while the other educators live in houses with children, both household sizes following the distributions outlined in the Parameterization section. The number of educator households is twice that required to fully supply the school due to the replacement process for symptomatic educators outlined in the Disease Progression section.

Initially, a proportion of all susceptible agents *R_init_* is marked as removed/recovered (*R*) to account for immunity caused by previous infection moving through the population. A single randomly chosen school attendee is chosen as a primary case and is made presymptomatic (*P*) to introduce a source of infection to the model. All simulations are run until there are no more potentially infectious (*E*, *P*, *I*, *A*) individuals left in the population and the institution is at full capacity. All results were averaged over 2000 trials.

### Estimating *β^H^*

Agents in the simulation were divided into two classes: “children” (ages 0 – 9) and “adults” (ages 25 – 44). Available data on contact rates^21^ was stratified into age categories of width 5 years starting at age 0 (0 – 5. 5 – 9, 10 – 14, etc.). The mean number of contacts per day 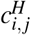 for each class we considered (shown in Eq. 2) was estimated by taking the mean of the contact rates of all age classes fitting within our presumed age ranges for children and adults.

For *β^H^* calibration, we created populations by generating a sufficient number of households to fill the institution in each of the three tested scenarios; 15: 2, 8: 2 and 7: 3. In each household, a single randomly chosen individual was infected (each member with equal probability) by assigning them a presymptomatic disease status *P*; all other members were marked as susceptible (disease status *S*). In each day of the simulation, each member of each household was allowed to interact with the infected member, becoming exposed to the disease with probability given in Eqn. 2. Upon exposure, they were assigned disease status *E*. At the beginning of each subsequent day, presymptomatic individuals proceeded to infected statuses *I* and *A*, and infected agents were allowed to recover as dictated by Fig. 1B and Supplementary Tab. S2. This cycle of interaction and recovery within each household was allowed to continue until all infected individuals were recovered from illness.

We did not allow exposed agents (status *E*) to progress to an infectious stage (*I* or *A*) since we were interested in finding out how many infections within the household would result *from a single infected household member*, as opposed to added secondary infections in later days. At the end of each trial, the specific probability of infection (*π_n_*) in each household *H_n_* was calculated by dividing the number of exposed agents in the household (*E_n_*) by the size of the household |*H_n_*| less 1 (accounting for the member initially infected). Single occupant households (|*H_n_*| = 1) were excluded from the calculation. The total probability of infection *π* was then taken as the mean of all *π_n_*, so that

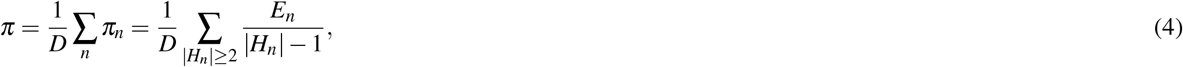

where *D* represents the total number of multiple occupancy households in the simulation. This modified disease simulation was run for 2000 trials each of different prospective values of *β^H^* ranging from 0 to 0.21. The means of all corresponding final estimates of the infection rate were taken per value of *β^H^*, and the value corresponding to a infection rate of 15% was interpolated.

## Data Availability

All data referred to in this manuscript are publicly available and the sources have been cited in the references list.

## Supplementary Appendix

### Sensitivity Analysis: varying *α*_0_ and *B_H_*

The parameter *β^H^* represents the rate of interaction in the household, and thereby regulates the spread of the disease. For each value of *α*_0_, increasing the rate of interaction in the home *β^H^* increases the number of infections produces for both RA (Supplementary Fig. S3) and ST (Supplementary Fig. S4) allocation. In most scenarios (7:3 RA being one of the exceptions), varying *α*_0_ (for constant *β^H^*) produces a small increase in the number of infections produced throughout the simulation. The rate of increase also depends on the number of children in the classroom; for the scenario 31:1 RA, increasing *β^H^* from 0.0545 to its baseline value 0.109 almost triples the number of total infections.subsection*Sensitivity Analysis - Varying *α*_0_ and *R_init_*

The parameter *R_init_* refers to the proportion of individuals we presume are recovered from some previous period of infection spread, while *α*_0_ is responsible for the rate of infection in common areas relative to the infection rate in the classroom. All other parameters are set to the baseline values given in Supplementary Tab. S2. These parameters were varied together by 50% in either direction. In Supplementary Figs. S5 and S6, increasing values of *R_init_* lower both the means and standard deviations of the total number of infections for each value of *α*_0_. Also, for each value of *R_init_*, the total number of infections produced increases with *α*_0_ . This shows opposing interaction between increasing common area infection and increasing initial recovery rate; one increases infection and the other lowers it (respectively).

### Sensitivity Analysis - Varying *α*_0_ and *λ_i_*

From Tab. S2, parameter *λ_i_* varies the amount of community infection in the model (infection due to other sources not modelled, such as public transport); be reminded that we assumed that the rate of community infection is effectively twice the baseline value for those individuals in the model not attending the school.

For each value of *α*_0_ in Supplementary Fig. S8, the total number of infections produced in the simulation increases with *λ* in each scenario with random allocation (RA), and also with grouping by household (ST, Supplementary Fig. S7). For each *λ*, there is no consistent relationship between the numbers of infections and the value of *α*_0_. This result is intuitive; though the effect is not pronounced, increasing the rate of community infection increases the total number of infections in each tested scenario.

### Supplementary Figures and Tables

**Table S1.** Times at which the mean proportions of presymptomatic (*P*), exposed (*E*), symptomatically infected (*I*) and asymptomatically infected (*A*) school attendees peak during the first 30 days of simulation with secondary spread with respect to each of the scenarios tested, and the corresponding peak number of cases.

| $\alpha_C$ | Status | Allocation | Peak Time | | | Maximum ($\times 10^{-4}$) | | |
| --- | --- | --- | --- | --- | --- | --- | --- | --- |
|  |  |  | 15:2 | 8:2 | 7:3 | 15:2 | 8:2 | 7:3 |
| 0.75 | P | RA | 12 | 0 | 0 | 304 | 200 | 200 |
|  |  | ST | 4 | 0 | 0 | 193 | 200 | 199 |
|  | E | RA | 18 | 3 | 3 | 497 | 252 | 204 |
|  |  | ST | 3 | 3 | 2 | 336 | 227 | 195 |
|  | I | RA | 12 | 2 | 2 | 49 | 37 | 35 |
|  |  | ST | 4 | 2 | 2 | 30 | 34 | 37 |
|  | A | RA | 19 | 5 | 5 | 165 | 198 | 111 |
|  |  | ST | 5 | 5 | 4 | 82 | 113 | 103 |
| $\alpha_C$ | Status | Allocation | 15:2 | 8:2 | 7:3 | 15:2 | 8:2 | 7:3 |
| 0.25 | P | RA | 0 | 0 | 0 | 118 | 200 | 200 |
|  |  | ST | 0 | 0 | 0 | 118 | 200 | 201 |
|  | E | RA | 4 | 3 | 5 | 96 | 113 | 128 |
|  |  | ST | 2 | 2 | 3 | 96 | 105 | 117 |
|  | I | RA | 2 | 2 | 2 | 19 | 27 | 21 |
|  |  | ST | 2 | 2 | 2 | 19 | 30 | 21 |
|  | A | RA | 5 | 4 | 5 | 69 | 111 | 100 |
|  |  | ST | 5 | 5 | 5 | 62 | 102 | 102 |

**Table S2.** Parameter definitions, baseline values and literature sources.

| Parameter | Meaning | Baseline Value | Source |
| --- | --- | --- | --- |
| $\eta$ | probability of symptomatic infection | 0.6 (adults)<br>0.4 (children) | TBD<br>TBD |
| $\delta$ | transition probability, $E \rightarrow P$ | 0.5/day | 35,36 |
| $\sigma$ | transition probability, $P \rightarrow I, A$ | 0.5/day | 35,36 |
| $\gamma_I$ | transition probability, $I \rightarrow R$ | 1.0/day | 35,36 |
| $\gamma_A$ | transition probability, $A \rightarrow R$ | 0.25/day | 35,36 |
| $c_{ij}^H$ | household contact matrix | ... | 21 |
| $\beta^H$ | transmission probability in households | 0.109 | 32, calibrated |
| $c_{ij}^C$ | room contact matrix | ... | 21 |
| $\beta^C$ | transmission probability in classrooms | $\beta^C = \alpha_C \beta^H$,<br>$\alpha_C = 0.75$ | 32, assumption |
| $\beta_{ij}^O$ | transmission probability in common areas | $\beta^O = \alpha_O \beta^C$,<br>$\alpha_O = 0.0025$ | 21,32, assumption |
| $\lambda_i$ | infection rate due to other sources | $1.16 \times 10^{-4}$/day | 33, estimated |
| $R_{init}$ | initial proportion with immunity | 0.1 | assumption |
| $\xi$ | probability of sibling attending same center | 0.8 | assumption |
| $o$ | proportion of childless educators | 0.36 | 31, assumption |
|  | household size distributions |  | 31 |

**Figure S1.**
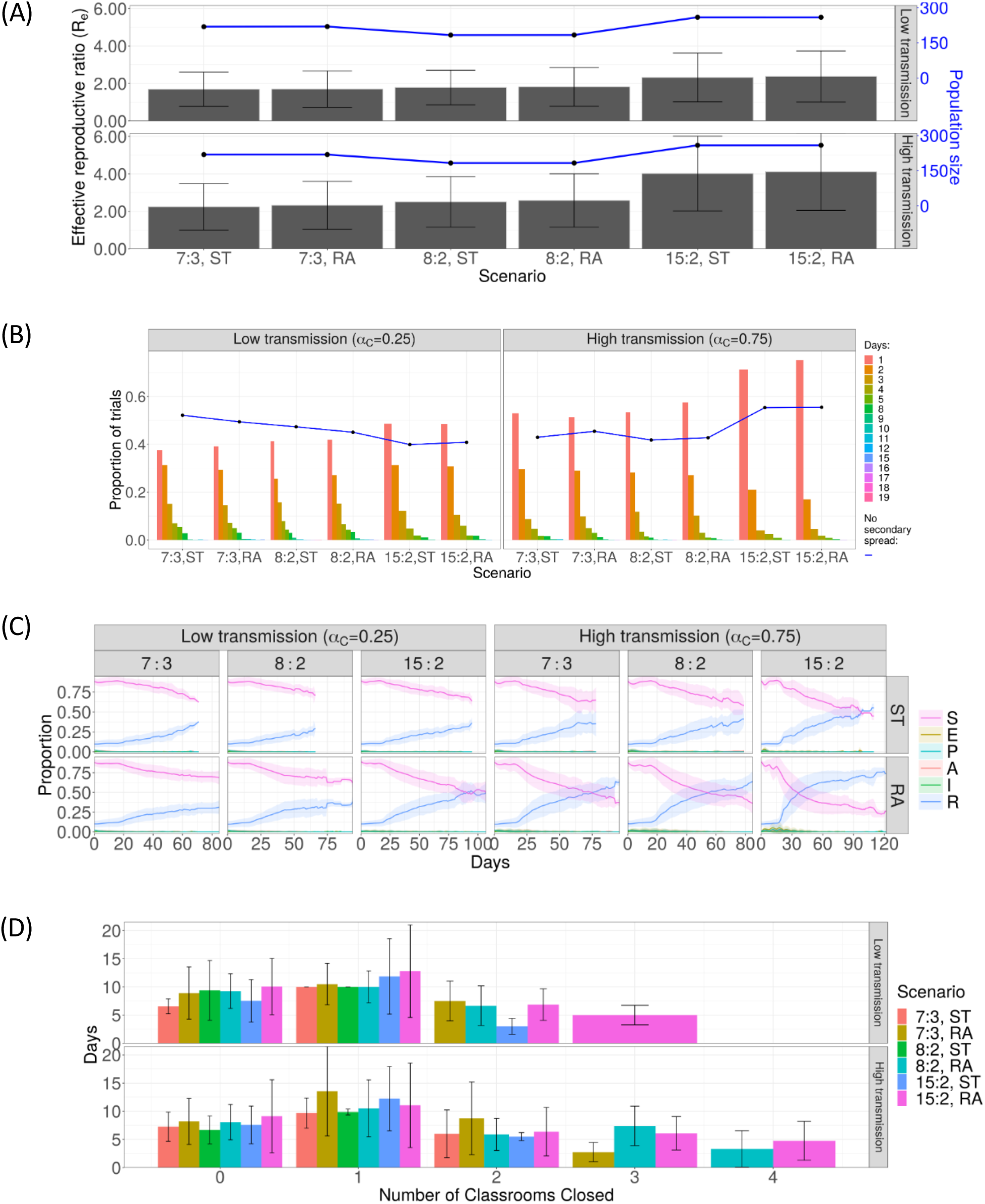
(fig:Combined2) Supplementary Results for Childcare Setting. (A) Bar chart showing the effective reproduction number *R_e_* in the entire population (with error bars denoting one standard deviation), with a line plot showing the mean population size. Both low and high transmission scenarios are shown. (B) Diagram showing the proportion of trials without secondary spread (curve), and the time taken to produce the first secondary infection (bar chart), both sorted by scenario. (C) Time series detailing the trends in the mean proportions of current school attendees in each stage of disease progression. Shaded ribbons around each curve show one standard deviation of the averaged time series. Only trials showing secondary spread were included in the ensemble means shown. (D) Bar chart showing the number of days for which some number of rooms in the school were closed due to disease outbreak. Scenarios are represented by different colours; the height of each bar gives the relevant ensemble mean with its standard deviation represented by error bars.

**Figure S2.**
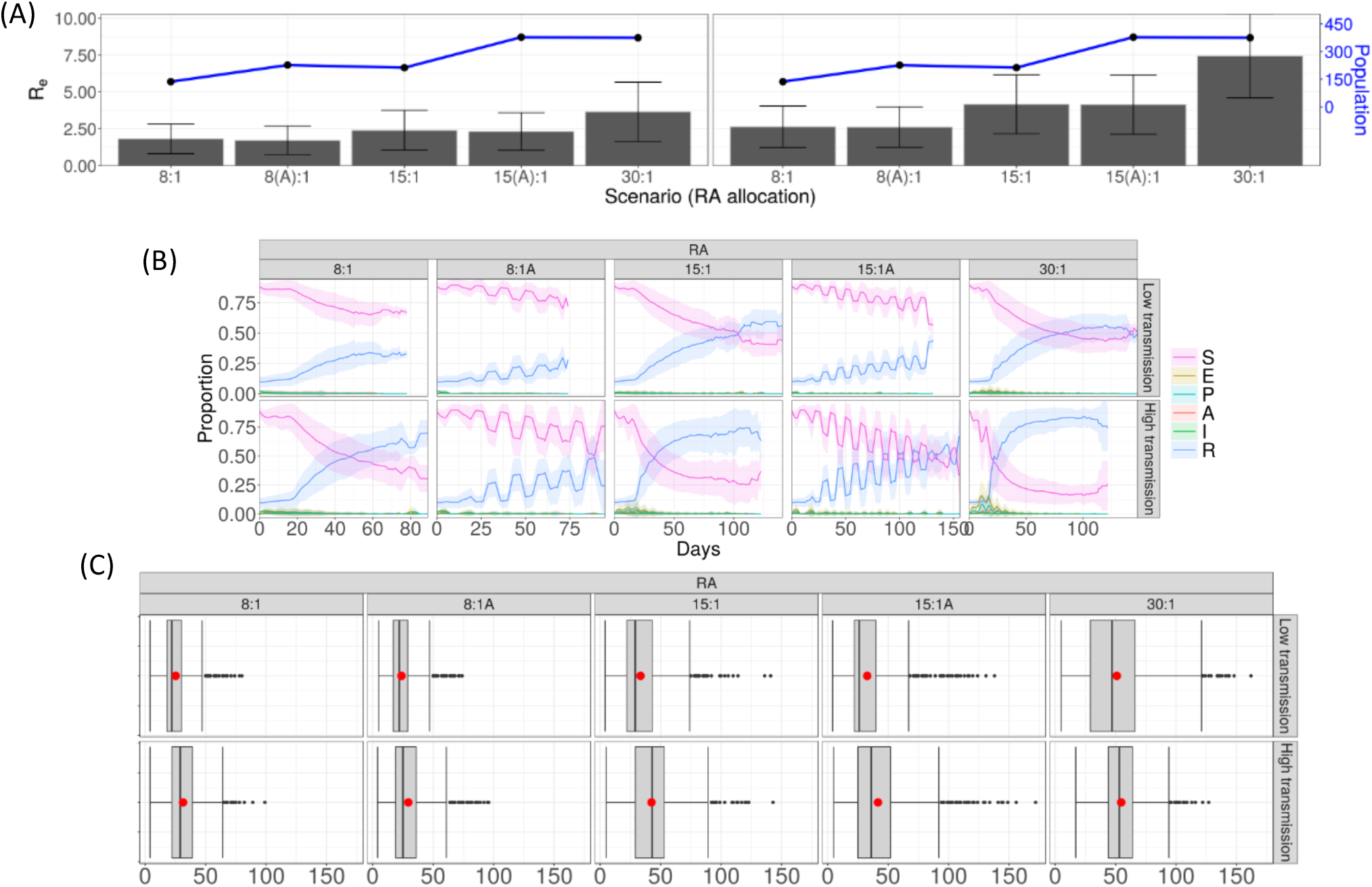
(fig:Combined1) Supplementary results for the primary school scenario. (A) Bar chart showing the effective reproduction number *R_e_* in the entire population (with error bars denoting one standard deviation), with a line plot showing the mean population size. Both low and high transmission scenarios are shown. (B) Time series showing the trends in the mean proportions of current school attendees in each stage of disease progression. Shaded ribbons around each curve show one standard deviation of the averaged time series. Only trials showing secondary disease spread were included in the ensemble means shown. (C) Box plots depicting the distribution of simulation durations for each scenario, describing the length of the outbreak. Red dots represent the arithmetic mean of the data.

**Figure S3.**
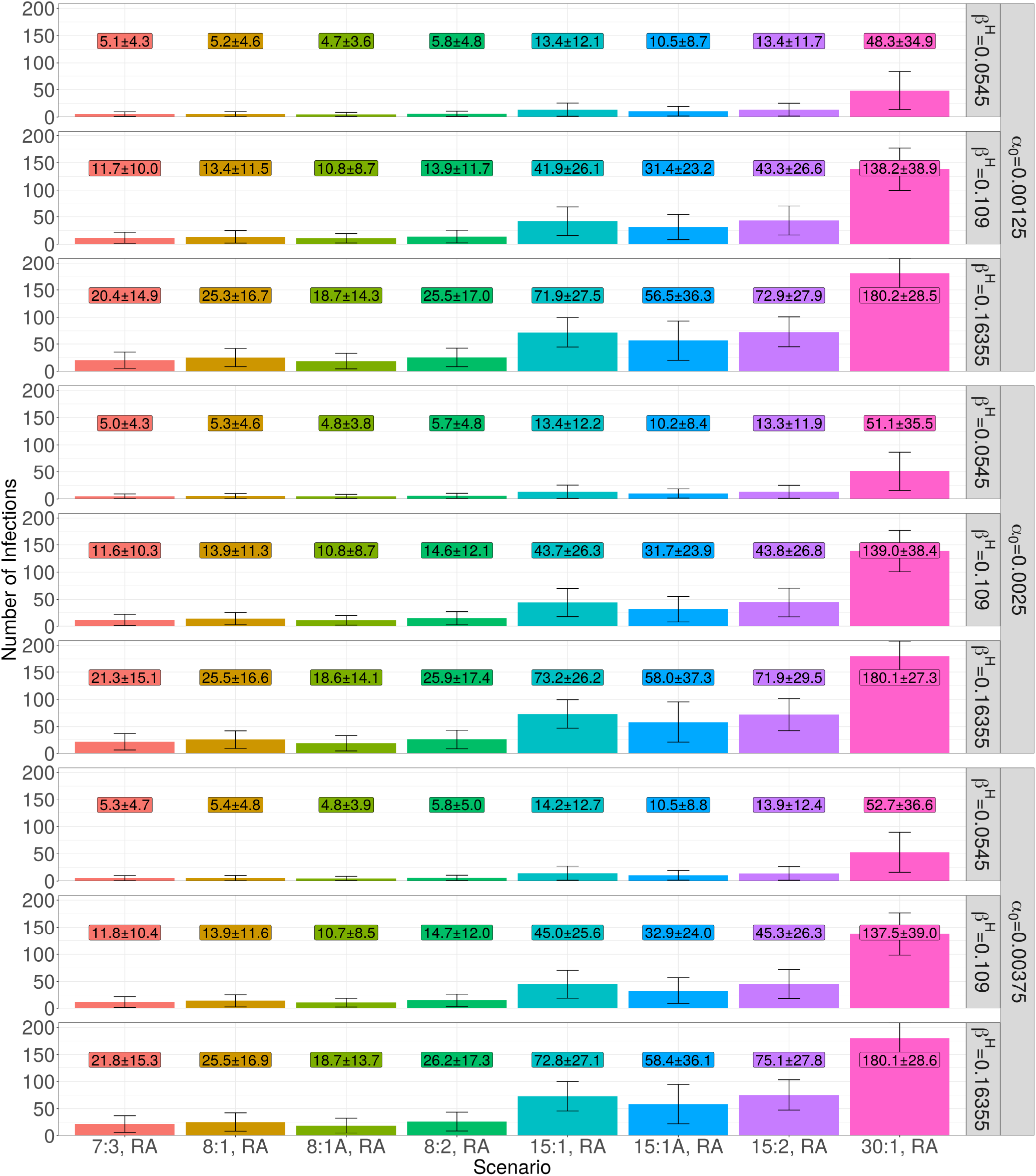
Results of varying the parameters *β^H^* and *α*_0_ by (50% each) on the total number of produced infections for RA allocation. Error bars denote a single standard deviation of the data used, and boxed text shows the corresponding mean and standard deviation.

**Figure S4.**
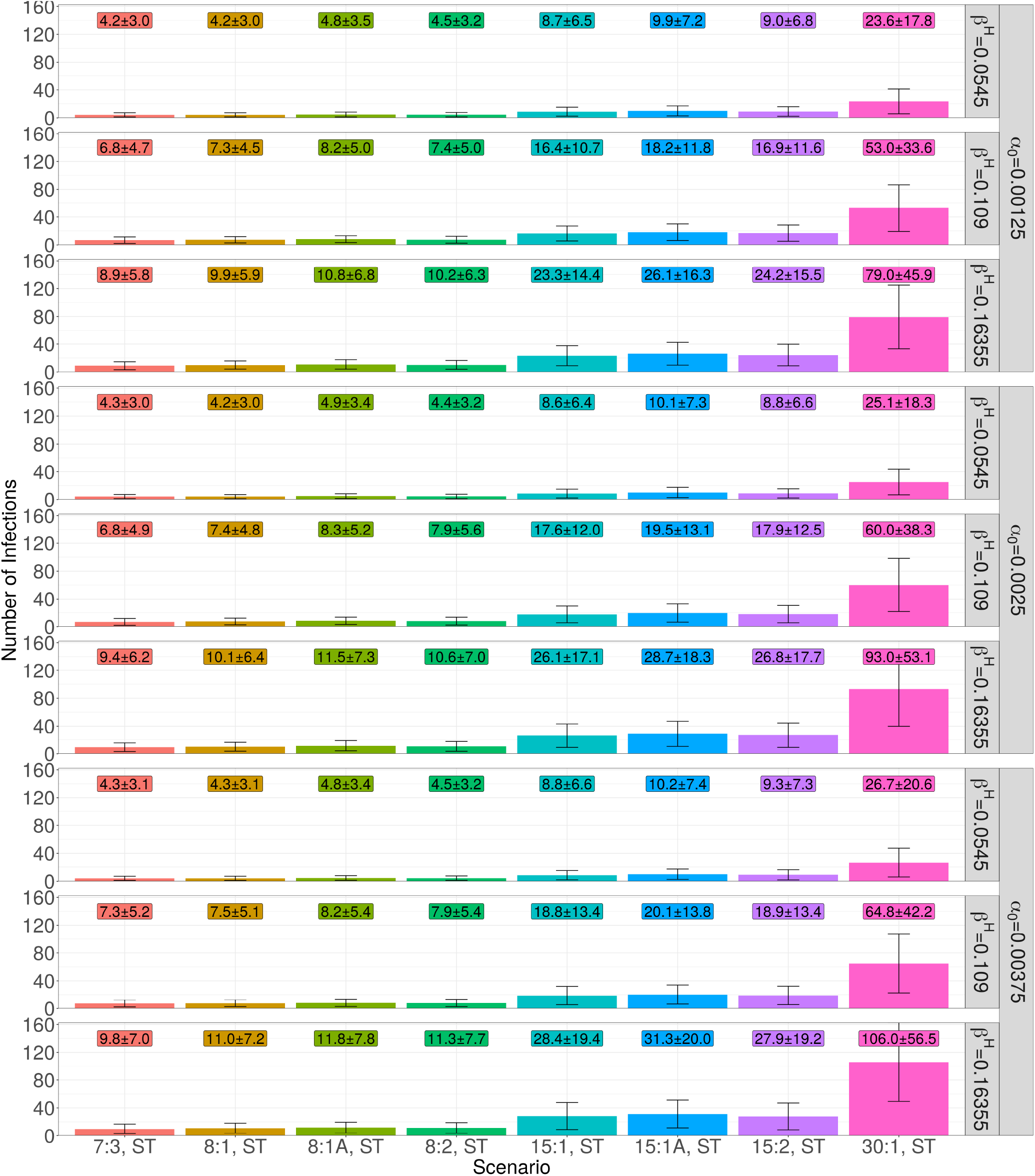
Results of varying the parameters *β^H^* and *α*_0_ by (50% each) on the total number of produced infections for ST allocation. Error bars denote a single standard deviation of the data used, and boxed text shows the corresponding mean and standard deviation.

**Figure S5.**
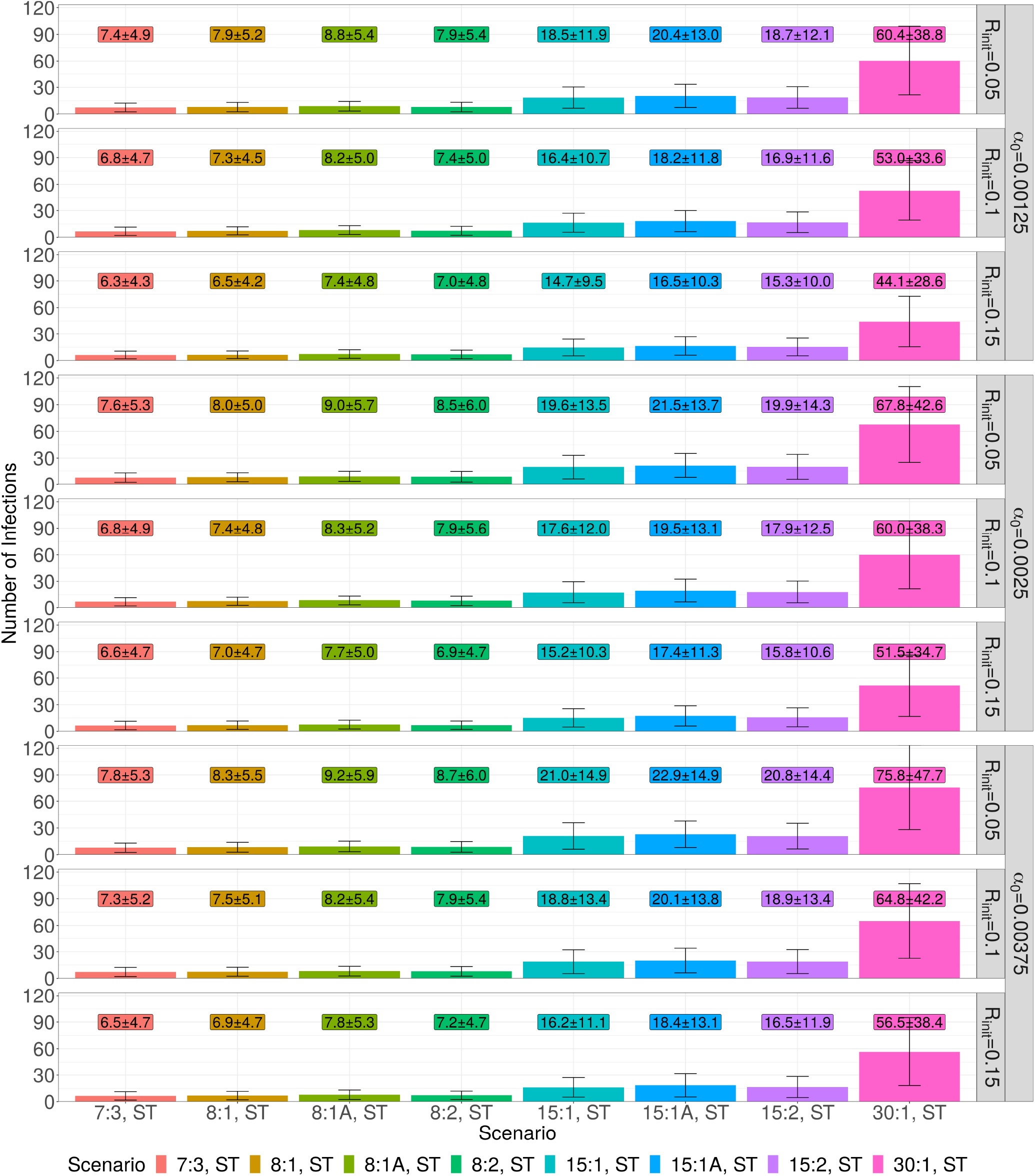
Results of varying the parameters *R_init_* and *α*_0_ by (50% each) on the total number of infections for ST allocation. Text in boxes denotes the mean and standard deviation of the data corresponding to the parameters and error bars denote a single standard deviation of the data used.

**Figure S6.**
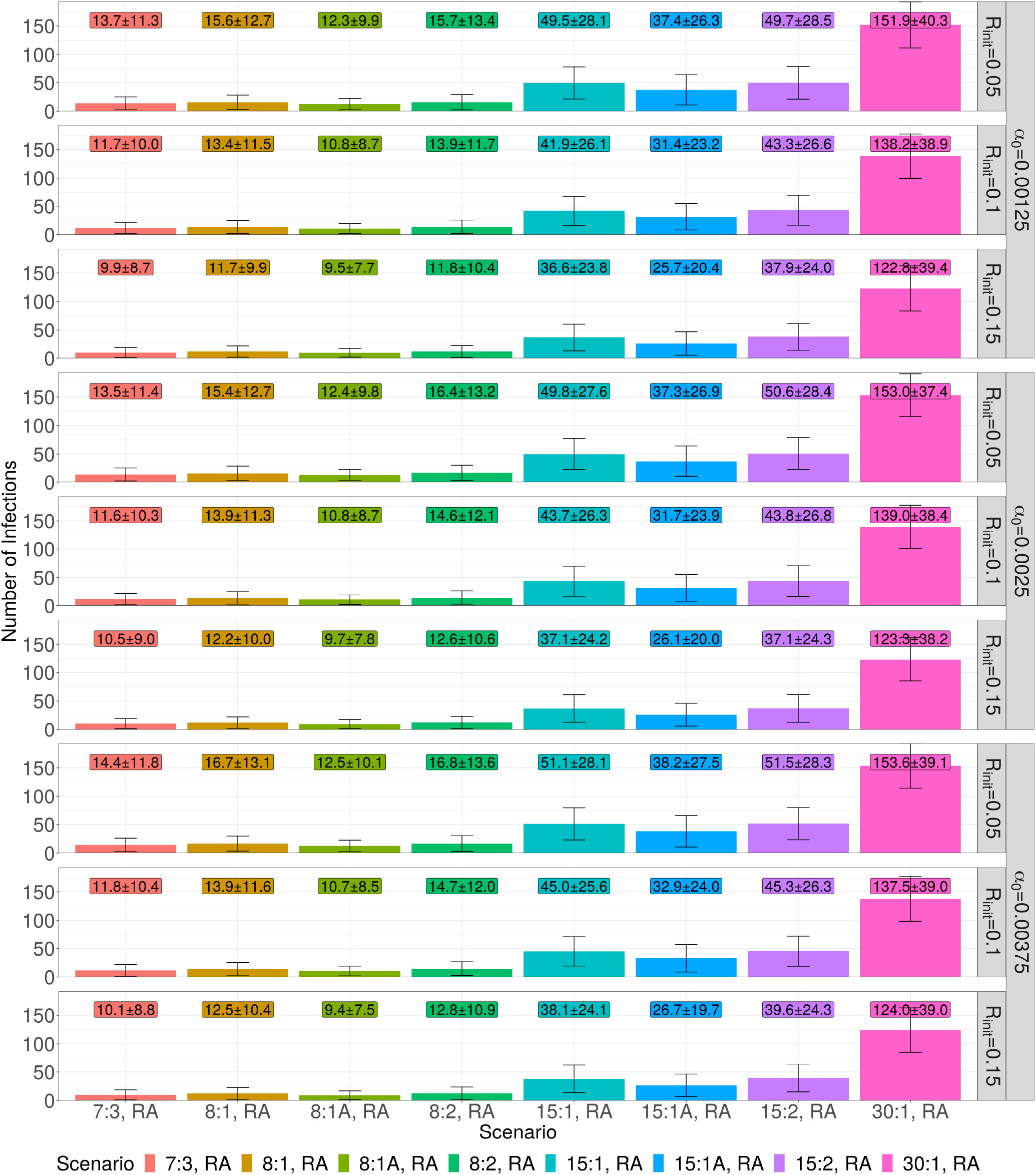
Results of varying the parameters *R_init_* and *α*_0_ by (50% each) on the total number of infections for RA allocation. Text in boxes denotes the mean and standard deviation of the data corresponding to the parameters and error bars denote a single standard deviation of the data used.

**Figure S7.**
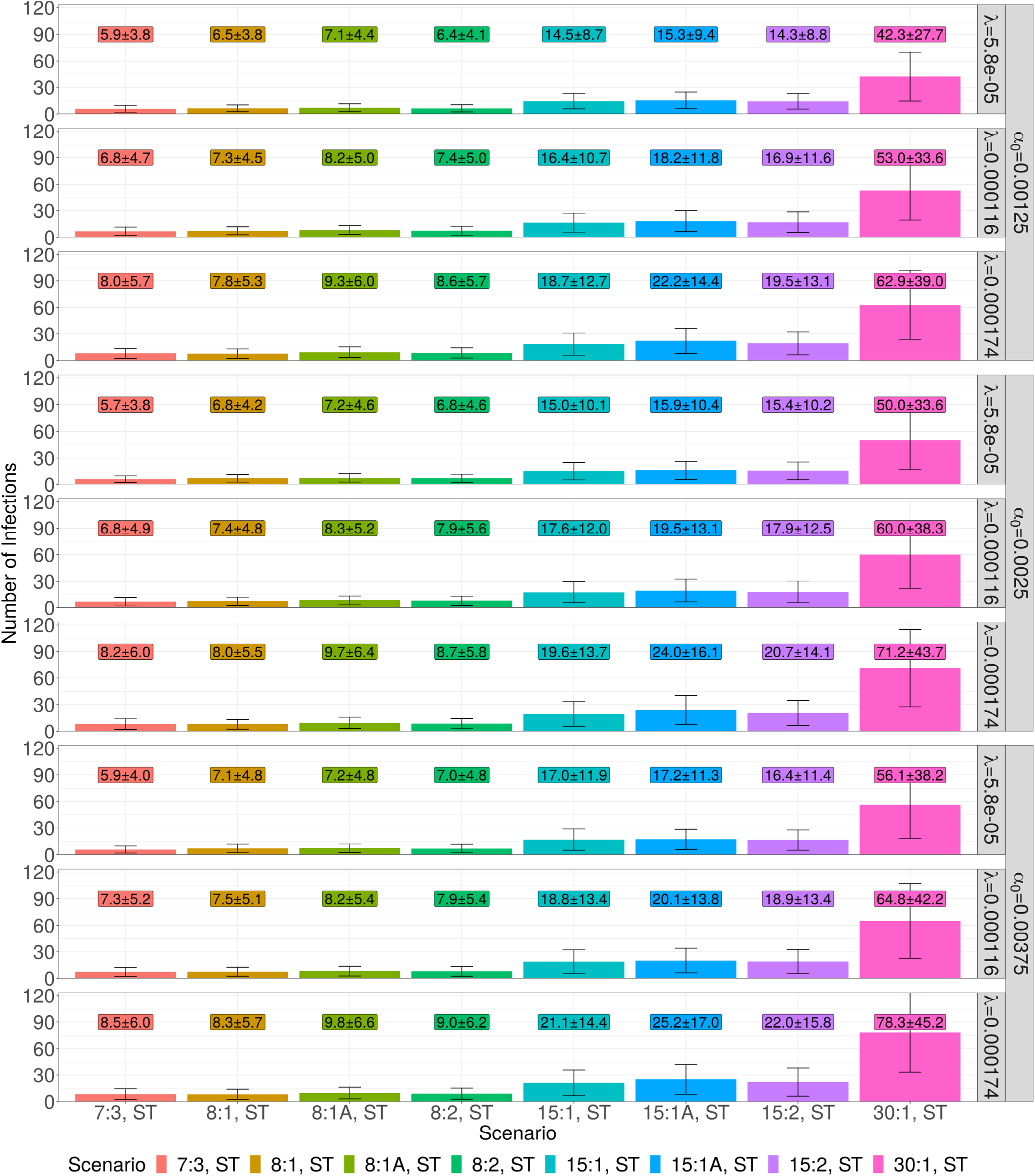
Results of varying the parameters *λ_i_* and *α*_0_ by (50% each) on the total number of infections for ST allocation. Text in boxes denotes the mean and standard deviation of the data corresponding to the parameters and error bars denote a single standard deviation of the data used.

**Figure S8.**
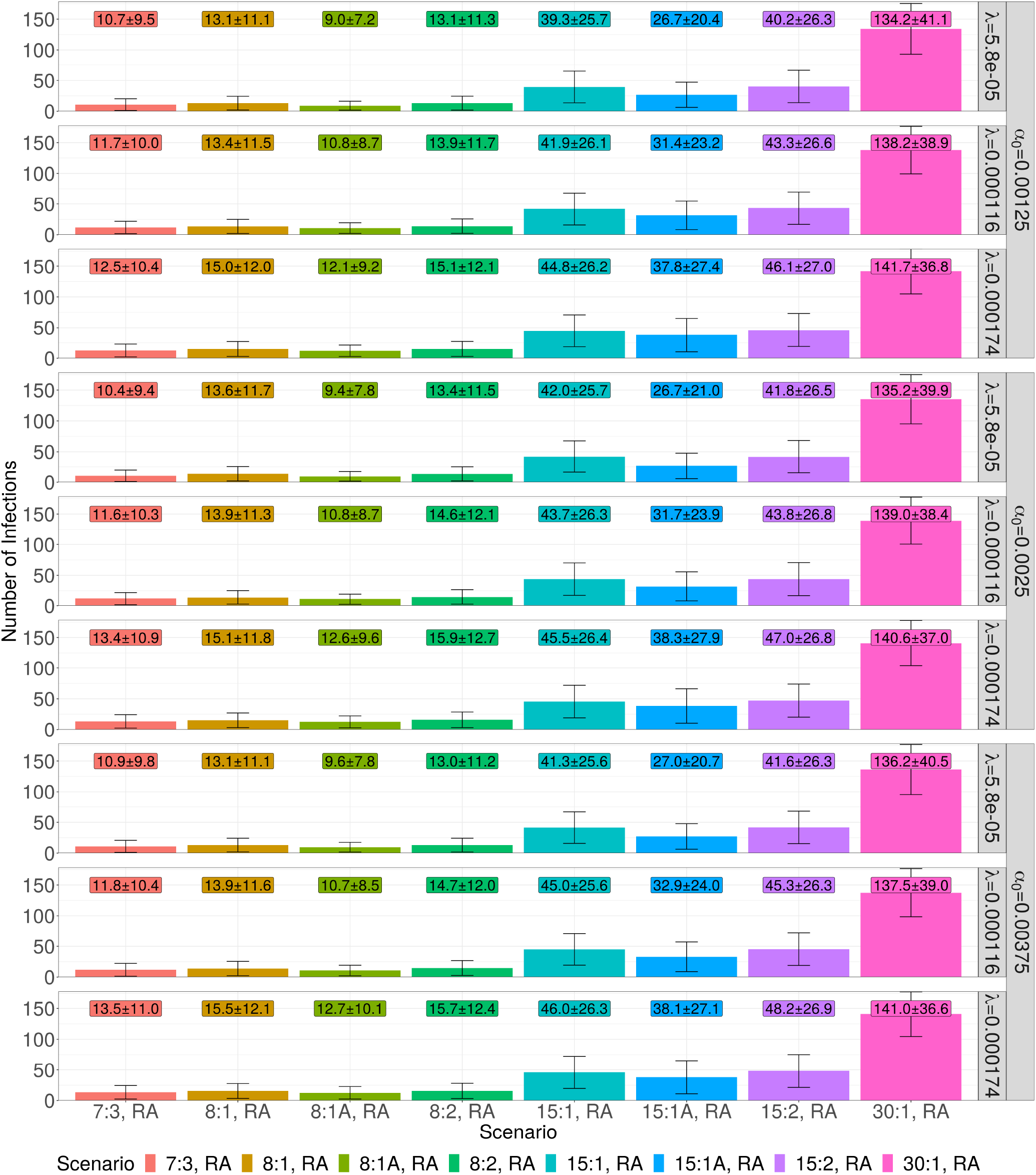
Results of varying the parameters *λ_i_* and *α*_0_ by (50% each) on the total number of infections for ST allocation. Text in boxes denotes the mean and standard deviation of the data corresponding to the parameters and error bars denote a single standard deviation of the data used.

